# From signal to knowledge: The diagnostic value of rawdata in artificial intelligence prediction of human data for the first time

**DOI:** 10.1101/2022.08.01.22278299

**Authors:** Bingxi He, Yu Guo, Yongbei Zhu, Lixia Tong, Boyu Kong, Kun Wang, Caixia Sun, Hailin Li, Feng Huang, Liwei Wu, Meng Wang, Fanyang Meng, Le Dou, Kai Sun, Tong Tong, Zhenyu Liu, Ziqi Wei, Wei Mu, Shuo Wang, Zhenchao Tang, Shuaitong Zhang, Jingwei Wei, Lizhi Shao, Mengjie Fang, Juntao Li, Shouping Zhu, Lili Zhou, Shuo Wang, Di Dong, Huimao Zhang, Jie Tian

## Abstract

Recently, image-based diagnostic technology has made encouraging and astonishing development. Modern medical care and imaging technology are increasingly inseparable. However, the current diagnosis pattern of Signal-to-Image-to-Knowledge inevitably leads to information distortion and noise introduction in the procedure of image reconstruction (Signal-to-Image). Artificial intelligence (AI) technologies that can mine knowledge from vast amounts of data offer opportunities to disrupt established workflows. In this prospective study, for the first time, we developed an AI-based Signal-to-Knowledge diagnostic scheme for lung nodule classification directly from the CT rawdata (the signal). We found that the rawdata achieved almost comparable performance with CT indicating that we can diagnose diseases without reconstructing images. Meanwhile, the introduction of rawdata could greatly promote the performance of CT, demonstrating that rawdata contains some diagnostic information that CT does not have. Our results break new ground and demonstrate the potential for direct Signal-to-Knowledge domain analysis.

## Introduction

The discovery of X-rays in 1895 ushered in a new era in the use of imaging for medical diagnostic purposes. Since then, the non-invasive medical imaging technology subverts the palpation and cut-and-see scheme [1]. The technological advances in medical imaging have been astounding over the past 120 years, and modern medical care is increasingly inseparable from imaging technology. Medical imaging is essential for humans, to allow clinicians to observe from the images and diagnose diseases. This process can be defined as a path of image-to-knowledge. However, recently, it is found that the human ability has become a bottleneck in this path hindering the accurate diagnosis and treatment of diseases [2][3].

The emergence of Artificial Intelligence (AI) technology partially solves the problem of the limited ability of humans in the diagnosis process [4][5][6][7]. AI could automatically mine the radiographic patterns that related to the occurrence and progression of diseases from the imaging data, and it has been shown to match and even surpass human abilities in many clinical applications [8][9][10][11][12]. The essential reason why AI could surpass humans may be that AI treats images as data rather than the visual image and extracts huge amounts of features for analysis [13][14]. However, the medical image is compressed or filtered data to fit the human eye, which may be insufficient or imperfect for diagnosis. Take computed tomography (CT) for example, the CT system first collects rawdata (signal) from the patient, then the reconstruction method converts rawdata to images (signal-to-image) [15]. Therefore, both AI-based and human-based diagnosis are processes of signal-to-image-to-knowledge. Medical images suffer from information distortion in both acquisition and reconstruction processes. The current high sampling frequency greatly compresses the influence of factors such as motion artifacts in the acquisition process, so the main reason for the loss of resolution is concentrated in the operations such as interpolation and sub-optimal statistical weighting in the reconstruction process [16]. Meanwhile, the unprocessed data size of rawdata is about 10 to 20 times larger than that of CT images (2GB compare with 180MB). The huge amount of information inside the rawdata is not optimally mined in current signal-to-image-to-knowledge process, and how to analysis rawdata is of great scientific interest.

Skipping the image process and going directly from signal to knowledge, will hopefully bring new breakthroughs in disease diagnosis. Inspired by this idea, several previous studies have talked about the potential value of analysis of rawdata [17][18][19], directly from signal to knowledge. De Man Q et.al. conducted a simulation experiment to detect and estimate the vessel centerline from rawdata in the sinogram domain [19]. They achieved encouraging initial results showing the feasibility of rawdata analysis for clinical CT analysis tasks. We have also reported our simulation results about lung cancer at American Association for Cancer Research (AACR) conference [20]. However, there is no study about signal-to-knowledge analysis in real clinical tasks of patients.

In this prospective study, for the first time, we developed an AI-based signal-to-knowledge diagnostic scheme for lung nodule classification directly from the CT rawdata (The flowchart was shown in **Fig. 1**). The value of rawdata alone (Discussion), as well as its added value to CT, are studied on 276 patients. We found that the rawdata achieved almost comparable performance with CT indicating that we can diagnose diseases without reconstructed images. Meanwhile, the introduction of rawdata could greatly promote the performance of CT, demonstrating that rawdata contains some diagnostic information that CT does not have. This research breaks the routinely used circle of image-based diagnosis, which may open up a new pathway of signal-to-knowledge for disease diagnosis.

**Figure 1.**
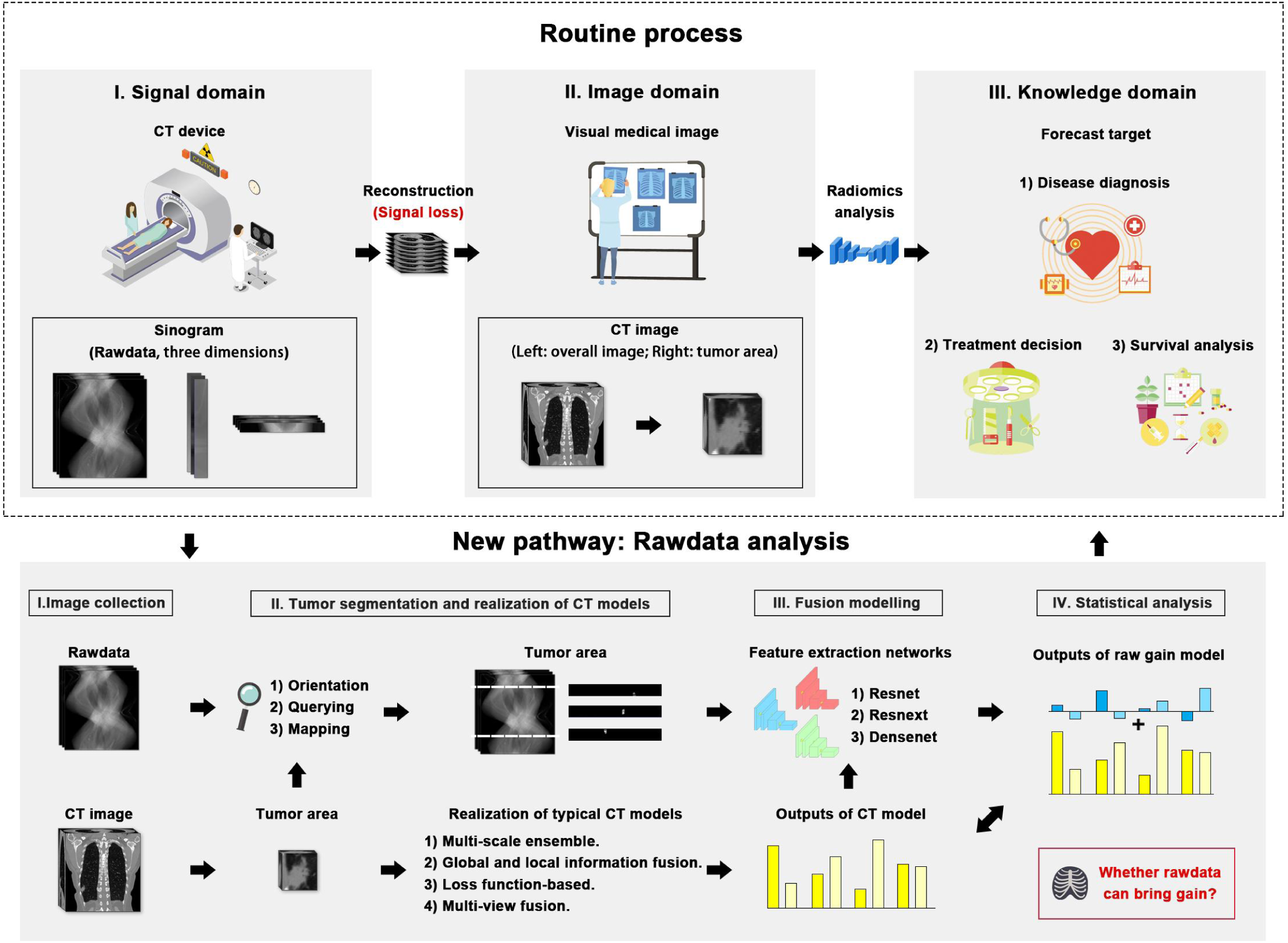
Flow chart of rawdata gain experiment.

## Results

### Clinical characteristics

The clinical characteristics was summarized in **Table S1**. A total of 276 patients were included and the number of patients in the training cohort, validation cohort and test cohort were 166, 55 and 55, respectively. Fifty percent (n = 138) patients were female and the mean of age in the entire dataset was 58.48 years. Furthermore, there were 21 (8%) small cell carcinoma, 35 (13%) squamous cell carcinomas and 149 (54%) adenocarcinomas. With respect to lesion location, most patients were identified as right upper lobe (n = 89, 32%), followed by left lower lobe (n = 67, 24%) and left upper lobe (n = 64, 23%) in all patients. For lung cancer diagnosis, most patients (n = 225, 82%) were evaluated as malignancy.

### Performance of CT model and rawdata gain model

This experiment explores the performance improvement that the residual fusion model (Methods) based on both rawdata and CT images can bring to the model based on CT images only. For further explore the repeatability and stability of this gain, we tested four different CT models (Abbreviation for CTM1∼CTM4; Methods) and adopted three backbone network architectures for rawdata feature extraction, namely Densenet121 (DN) [21], Resnet18 (RE) [22] and Resnext18 (RX) [23]. For each CT model (CTM), three rawdata gain models (RGM) based on different backbone feature extraction networks were constructed. The performance of each RGM was compared with the original CTM. The receiver operating characteristic curves (ROC) and its area under the curve (AUC) of four CTMs and the corresponding RGMs based on difference backbone networks is shown in **Fig. 2**.

**Figure 2.**
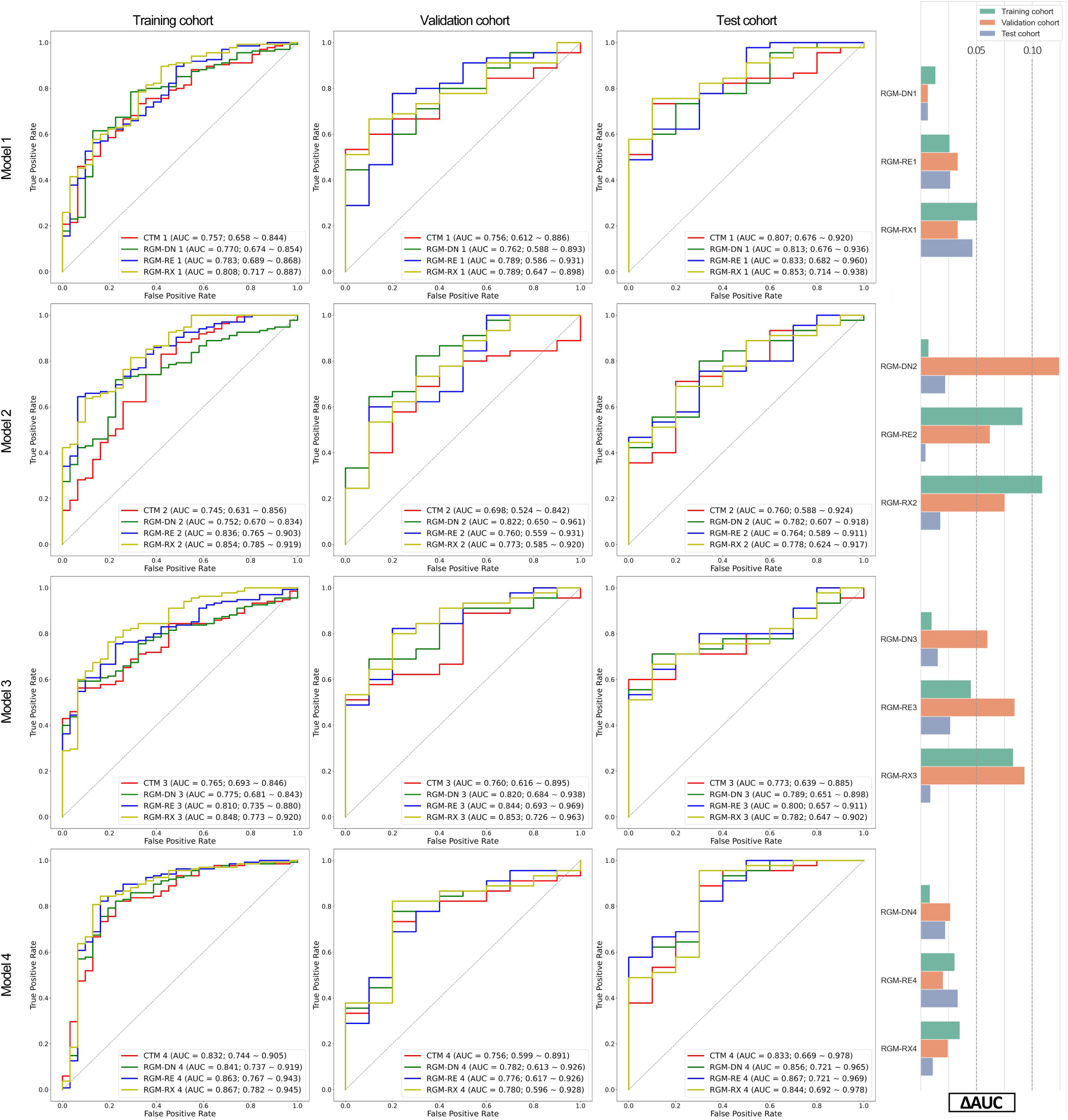
CT model and raw data gain results. (A) shows the ROC curves of each CTM and residual fusion model based on different backbone feature extraction networks; (B) Bar graph of reaction AUC gain. DM, Densenet121; RE, Resnet18, RX, Renext18.

For each CTM, the residual fusion models based on different backbone networks can obtain better classification performance on training, validation and test cohorts. For CTM1 model, the fusion model that produced the maximum performance improvement for the training cohort is RGM-RX1, and its AUC improvement can reach 0.051 (from 0.757 to 0.808). The fusion model that produced the maximum performance improvement for the validation cohort is RGM-RE1 and RGM-RX1, and its AUC improvement can reach 0.033 (from 0.756 to 0.789). The fusion model that produced the maximum performance improvement for the test cohort is RGM-RE1, and its AUC improvement can reach 0.046 (from 0.807 to 0.853). For CTM2 model, the fusion model that produced the maximum performance improvement for the training cohort is RGM-RX2, and its AUC improvement can reach 0.109 (from 0.745 to 0.854). The fusion model that produced the maximum performance improvement for the validation cohort is RGM-DM2, and its AUC improvement can reach 0.124 (from 0.698 to 0.822). The fusion model that produced the maximum performance improvement for the test cohort is RGM-DM2, and its AUC improvement can reach 0.022 (from 0.760 to 0.782). For CTM3 model, the fusion model that produced the maximum performance improvement for the training cohort is RGM-RX3, and its AUC improvement can reach 0.083 (from 0.765 to 0.848). The fusion model that produced the maximum performance improvement for the validation cohort is RGM-RX3, and its AUC improvement can reach 0.093 (from 0.760 to 0.853). The fusion model that produced the maximum performance improvement for the test cohort is RGM-RE3, and its AUC improvement can reach 0.027 (from 0.773 to 0.800). For CTM4 model, the fusion model that produced the maximum performance improvement for the training cohort is RGM-RX4, and its AUC improvement can reach 0.035 (from 0.832 to 0.867). The fusion model that produced the maximum performance improvement for the validation cohort is RGM-DM4, and its AUC improvement can reach 0.026 (from 0.756 to 0.782). The fusion model that produced the maximum performance improvement for the test cohort is RGM-RE4, and its AUC improvement can reach 0.034 (from 0.833 to 0.867). Overall, using Resnext18 as the backbone network of rawdata feature extraction can obtain the maximum average performance improvement on the three cohorts.

### Image feature distribution of the RGMs and gain stability analysis

We performed t-SNE dimensionality reduction on all deep learning features obtained by different feature extraction networks and counted the true positives, false positives, true negatives and false negatives of each patient (**Table 1**). Besides, we assigned different colors and markers to visualize them in the same coordinate system (**Fig. 3**). It can be observed from the **Fig. 3** that the results of the various RGMs within each CTM are relatively similar, even though they use different feature extraction networks. **Table 1** also shows the same situation. The gain of the RGMS inside each CTM is approaching the same trend, such as improving malignant or benign detectable rates. Meanwhile, RGM-RE1, RGM-DN2, RGM-RE2, RGM-DN3, RGM-DN4, RGM-RE4, and RGM-RX4 could achieve a significant increase in the detectable rate of one category at the expense of a small number of the other category detectable rates.

**Table 1.**
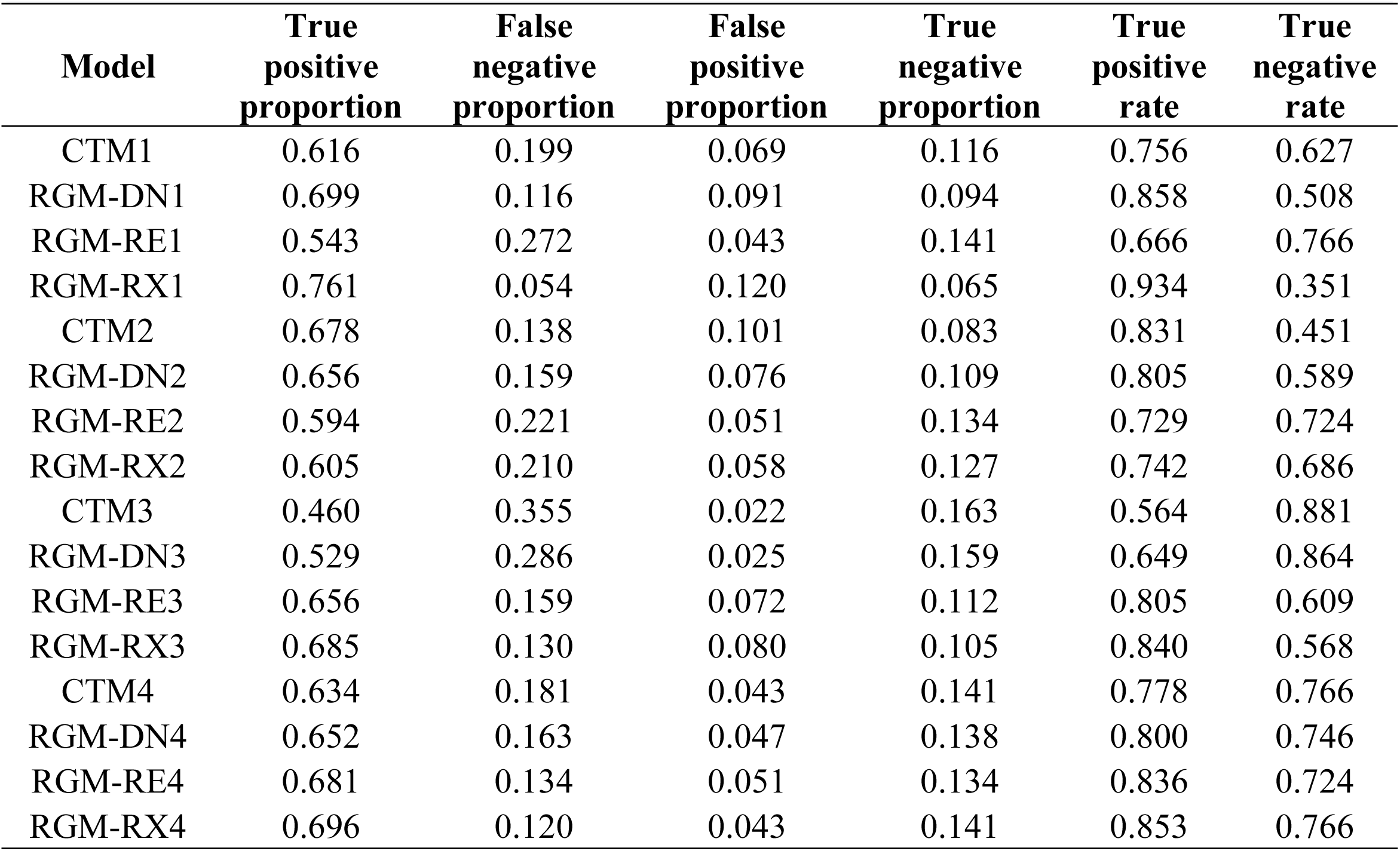
Detailed result statistics of raw data for CT models

**Figure 3.**
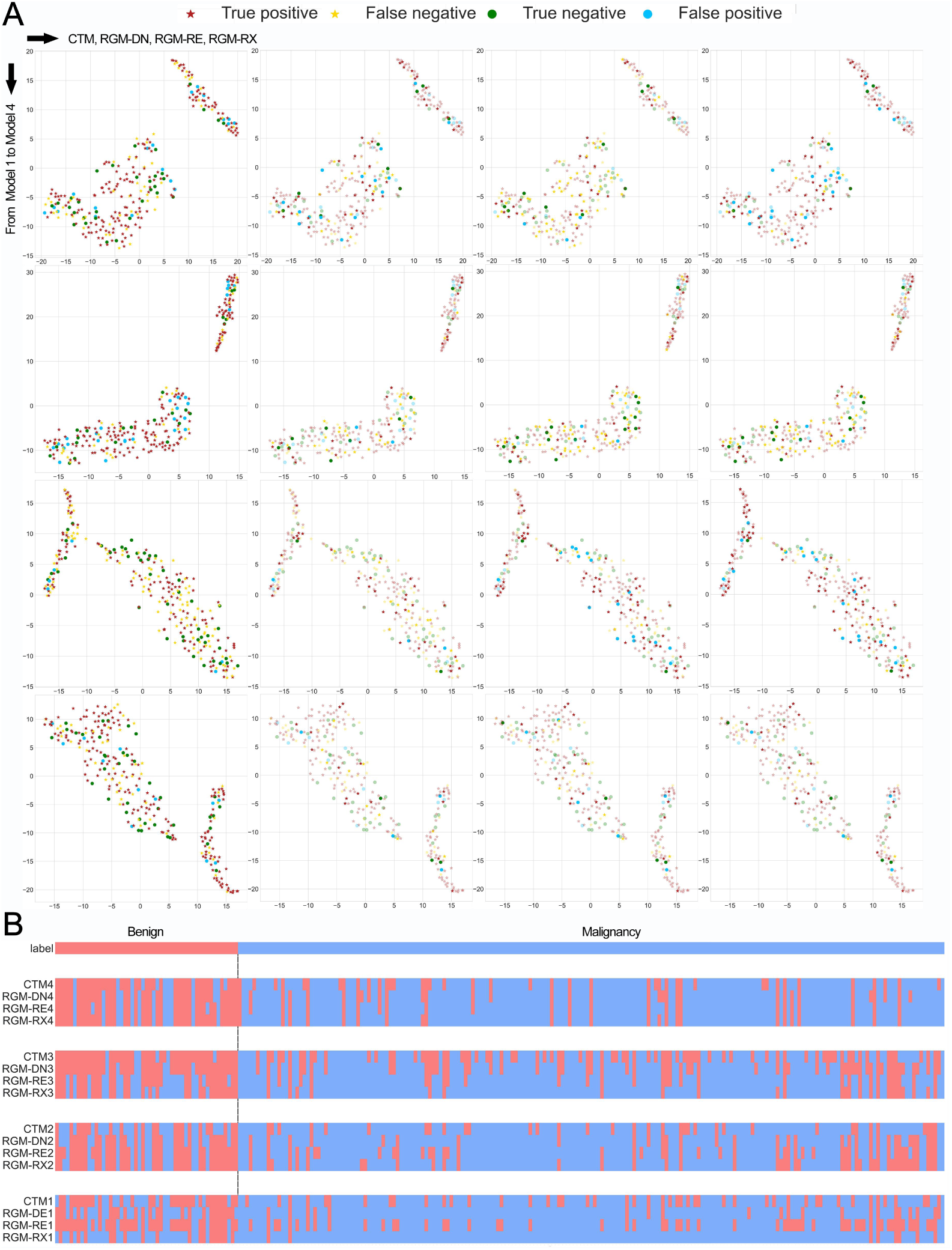
Feature distribution of the RGMs and the heatmap of prediction results. (A) shows the feature distribution in the RGM, and different marks and colors reflect different authenticity and prediction categories. (B) shows the prediction results of all real categories, CTMs and RGMs.

Therefore, we calculated the optimization rate and error rate of each RGM for the CTM, and also calculated the proportion of at least 2 model optimizations to all optimization samples, which can reflect the stability of rawdata’s gain. All results were summarized in **Table S2**.

The results show that the analysis method incorporating the rawdata has a high optimization rate for the CTM 1∼3 and is greater than the error rate, which is also reflected in the improvement of AUC. In addition, although different feature extraction networks were used to analysis the rawdata, the proportion of at least two networks that can be optimized in each CTM is about 80%. Finally, we found that 7 samples were mispredicted within 4 CTMs. For these 7 samples, the income of rawdata can correct the prediction results of the 6 CTMs, and the corrected model exists in each CTM. In summary, the gain of the rawdata for the CTM is very stable.

### Visual statistics and analysis of the RGMs

To better explain the prediction process of RGM, we visualized the region of most interest in the RGM by using Gradient-weighted Class Activation Mapping (Grad-CAM). The predictive results of RGM were most dependent on the information of the RGM-discovered suspicious areas. **Fig. 4** illustrated the lesion masks and corresponding attention maps from different views of the rawdata. From **Fig. 4**, we can see that the RGMs can always focus on areas of lesion for prediction although the input data includes some non-lesion areas. We also calculated the average attention score of each voxel in lesion and non-lesion in rawdata (Method), and the result showed that the attention score of the lesion area was 1-2 times as high as the non-lesion area.

**Figure 4.**
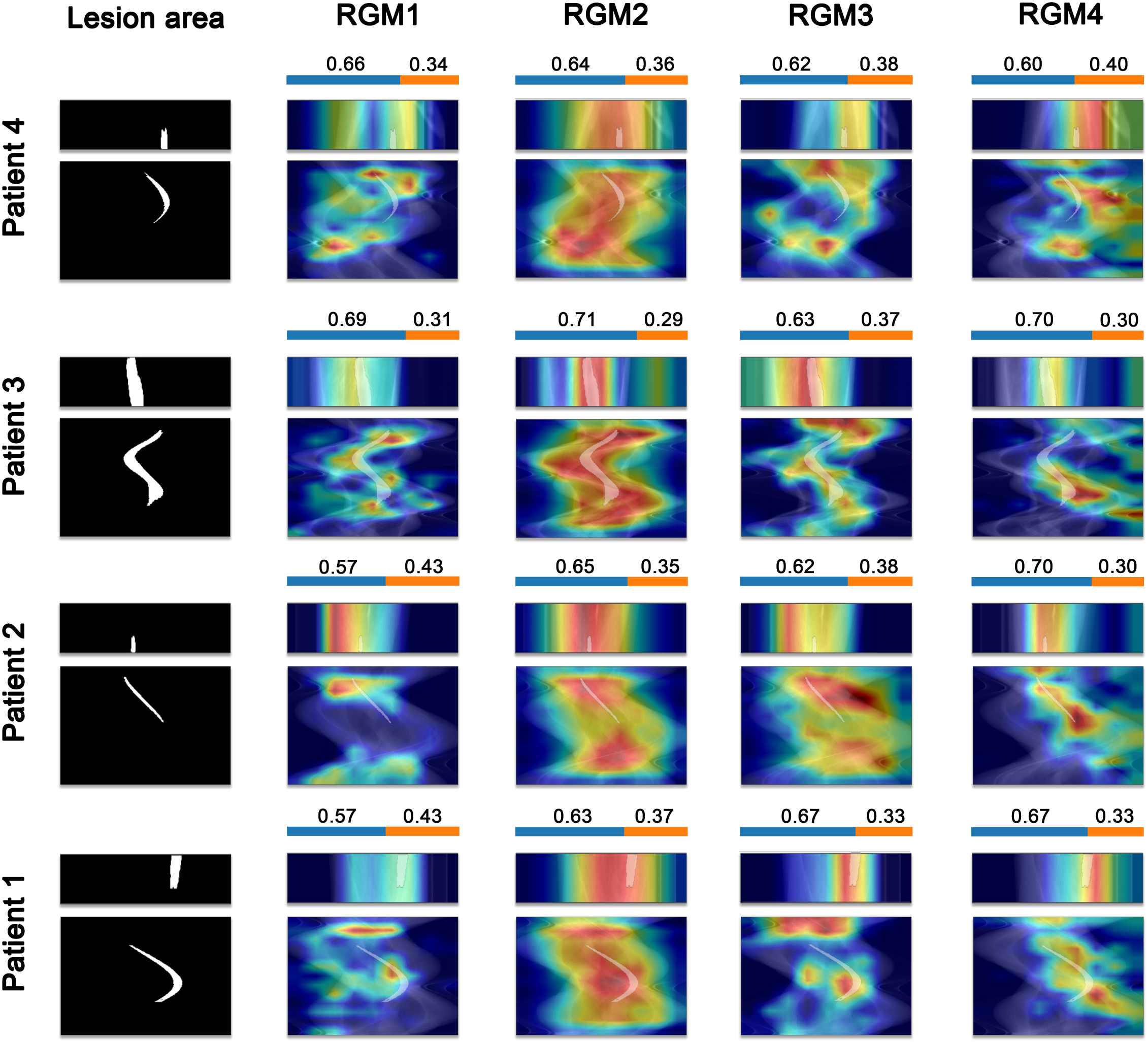
Lesion trajectory in rawdata and the Grad-Cam graphs of the RGMs.

### Stratified analysis of different malignant subgroups

The results of the subgroup analysis for age, sex and lesion size were shown in the **Table S3** and **Table 2**.

**Table 2.** The performance of 12 raw gain models in subgroup analysis. S

| AUC<br>95%CI |  | RGM-DN 1 | RGM-RE 1 | RGM-RX 1 | RGM-DN 2 | RGM-RE 2 | RGM-RX 2 | RGM-DN 3 | RGM-RE 3 | RGM-RX 3 | RGM-DN 4 | RGM-RE 4 | RGM-RX 4 |
| --- | --- | --- | --- | --- | --- | --- | --- | --- | --- | --- | --- | --- | --- |
| Age | <=60 | 0.731<br>(0.647-0.825) | 0.760<br>(0.667-0.844) | 0.756<br>(0.667-0.837) | 0.764<br>(0.668-0.845) | 0.790<br>(0.709-0.874) | 0.786<br>(0.705-0.867) | 0.800<br>(0.719-0.870) | 0.825<br>(0.743-0.895) | 0.824<br>(0.750-0.894) | 0.829<br>(0.749-0.903) | 0.836<br>(0.746-0.915) | **0.837**<br>**(0.746-**<br>**0.924)** |
|  | >60 | 0.801<br>(0.680-0.908) | 0.781<br>(0.664-0.894) | 0.804<br>(0.700-0.900) | 0.740<br>(0.610-0.859) | 0.792<br>(0.678-0.901) | 0.815<br>(0.699-0.932) | 0.744<br>(0.636-0.833) | 0.767<br>(0.648-0.863) | 0.799<br>(0.673-0.914) | 0.808<br>(0.661-0.926) | 0.835<br>(0.690-0.945) | **0.845**<br>**(0.713-**<br>**0.949)** |
| Sex | Male | 0.693<br>(0.584-0.794) | 0.698<br>(0.591-0.793) | 0.697<br>(0.601-0.792) | 0.737<br>(0.626-0.826) | 0.725<br>(0.608-0.837) | 0.714<br>(0.600-0.824) | 0.702<br>(0.605-0.791) | 0.736<br>(0.634-0.834) | 0.717<br>(0.617-0.822) | 0.793<br>(0.695-0.877) | 0.791<br>(0.686-0.889) | **0.810**<br>**(0.707-**<br>**0.897)** |
|  | Female | 0.808<br>(0.716-0.898) | 0.825<br>(0.728-0.901) | 0.835<br>(0.749-0.901) | 0.786<br>(0.695-0.871) | 0.842<br>(0.749-0.916) | 0.859<br>(0.777-0.927) | 0.852<br>(0.780-0.921) | 0.863<br>(0.784-0.920) | **0.885**<br>**(0.818-**<br>**0.945)** | 0.849<br>(0.741-0.944) | 0.873<br>(0.764-0.958) | 0.863<br>(0.739-0.954) |
| Tumor<br>Size | <=23mm | 0.771<br>(0.677-0.849) | 0.786<br>(0.689-0.869) | 0.815<br>(0.732-0.893) | 0.775<br>(0.685-0.863) | 0.821<br>(0.743-0.894) | 0.828<br>(0.752-0.901) | 0.800<br>(0.722-0.869) | 0.823<br>(0.736-0.888) | **0.847**<br>**(0.781-**<br>**0.916)** | 0.821<br>(0.729-0.906) | 0.836<br>(0.744-0.925) | 0.839<br>(0.731-0.926) |
|  | >23mm | 0.737<br>(0.603-0.847) | 0.746<br>(0.627-0.854) | 0.709<br>(0.582-0.815) | 0.748<br>(0.639-0.847) | 0.753<br>(0.634-0.864) | 0.754<br>(0.641-0.859) | 0.759<br>(0.657-0.850) | 0.782<br>(0.684-0.874) | 0.775<br>(0.666-0.868) | 0.822<br>(0.712-0.924) | **0.833**<br>**(0.703-**<br>**0.925)** | 0.831<br>(0.729-0.928) |
| Tumor<br>Location | superior<br>left | 0.679<br>(0.520-0.836) | 0.667<br>(0.464-0.836) | 0.743<br>(0.616-0.881) | 0.638<br>(0.469-0.812) | 0.683<br>(0.480-0.847) | 0.737<br>(0.517-0.899) | 0.824<br>(0.702-0.925) | 0.812<br>(0.701-0.916) | 0.810<br>(0.680-0.914) | 0.861<br>(0.754-0.953) | 0.885<br>(0.773-0.964) | **0.909**<br>**(0.829-**<br>**0.979)** |
|  | inferior<br>left | 0.796<br>(0.638-0.915) | 0.789<br>(0.652-0.921) | 0.769<br>(0.600-0.896) | 0.741<br>(0.584-0.881) | 0.759<br>(0.607-0.905) | 0.771<br>(0.599-0.911) | 0.775<br>(0.633-0.894) | 0.793<br>(0.656-0.907) | 0.812<br>(0.650-0.934) | 0.813<br>(0.651-0.953) | **0.840**<br>**(0.688-**<br>**0.969)** | 0.833<br>(0.661-0.961) |
|  | superior<br>right | 0.739<br>(0.618-0.855) | 0.814<br>(0.698-0.912) | 0.785<br>(0.673-0.876) | 0.810<br>(0.704-0.900) | 0.843<br>(0.740-0.930) | 0.811<br>(0.685-0.901) | 0.756<br>(0.633-0.864) | 0.802<br>(0.675-0.909) | 0.815<br>(0.675-0.899) | 0.843<br>(0.730-0.936) | 0.848<br>(0.741-0.934) | **0.849**<br>**(0.729-**<br>**0.939)** |
|  | Inferior<br>right | 0.821<br>(0.638-0.957) | 0.809<br>(0.655-0.957) | 0.795<br>(0.617-0.937) | **0.872**<br>**(0.744-**<br>**0.970)** | 0.849<br>(0.701-0.970) | 0.849<br>(0.700-0.970) | 0.786<br>(0.617-0.923) | 0.835<br>(0.705-0.961) | 0.812<br>(0.616-0.959) | 0.721<br>(0.430-0.942) | 0.764<br>(0.470-0.981) | 0.775<br>(0.524-0.962) |

In the subgroup with age of <= 60, RGM-RX 4 and CTM 4 achieved the similar highest model performance, with AUC of 0.837 (0.746-0.924) and 0.831 (0.749-0.904), respectively; In the subgroup with age of > 60, RGM-RX 4 achieved the highest model performance with an AUC of 0.845 (0.713-0.949), which outperformed the best CTM (CTM4 with an AUC of 0.790). In the male subgroup, CTM4 and RGM-RX 4 performed best, with a similar AUC of 0.804 (0.706-0.882) and 0.810 (0.707-0.897), respectively; In the female subgroup, RGM-RX 3 achieved the highest performance with an AUC of 0.885 (0.818-0.945), far exceeding the best CTM (CTM4 with an AUC of 0.823 (0.720-0.920)). In the subgroup with lesion size <=23mm, RGM-RX 3 achieved the highest model performance with an AUC of 0.847 (0.781-0.916), far exceeding the best CTM (CTM4 with an AUC of 0.806); In the subgroup of lesion size >23mm, CTM 4 and RGM-RE 4 showed the similar highest model performance, with AUC of 0.819 (0.719-0.906) and 0.833 (0.703-0.925), respectively. As for the lesion location subgroups, in addition to the similar performance of CTM 4 and RGM-RX 4 in the subgroup of superior lobe of left lung, RGM outperformed the CTM, with AUC of 0.840 vs. 0.812 in the subgroup of inferior lobe of left lung, 0.849 vs. 0.807 in the subgroup of superior lobe of right lung, and 0.872 vs. 0.843 in the subgroup of inferior lobe of right lung.

## Discussion

In this prospective study, for the first time, we validated the potential value of rawdata in real clinical practice. Interestingly, the rawdata analysis showed comparable performance with CT images, which indicates that leveraging non-image information holds promise as an alternative to image-based methods. Moreover, the add value of rawdata to CT images was also confirmed in this study, which means that the combination of non-image and image data will further promote the advance of disease diagnosis. This study proposed and validated a feasible method for diagnosis without image reconstruction, and it has the potential to change existing imaging-based diagnosis and treatment strategies.

The classification of benign and malignant pulmonary nodules is a matter of great clinical concern[24][25][26]. This study explores the feasibility of rawdata analysis in classifying indeterminate lung nodules greater than 2 cm in size. The results indicated that rawdata can well discriminate malignant nodules from benign nodules. Meanwhile, the AUCs of the rawdata in the training cohort, validation cohort and test cohort are 0.768 (95%CI: 0.681∼0.851), 0.760 (95%CI: 0.558∼0.922) and 0.782 (95%CI: 0.592∼0.924), respectively (**Extended Data Fig. 1**), and there is no statistical difference between the performances of rawdata and CT, which means that the classification of lung nodules may not need image reconstruction and clinician participation. Think further, the rawdata model could be applied to the majority of grassroots hospitals, who have mainstream CT systems but lack technical personnel and clinicians.

Our study showed that the introduction of rawdata to CT had an overall improvement over different CTMs, no matter which backbone network was used. This indicates that rawdata has unique information which may be lost during the reconstruction processing. Moreover, the compared with CTM, RGMs showed better stability on the training cohort, validation cohort, and test cohort. The combination of both non-image and image data could make the model robust. In addition, we also performed intra-CT and inter-CT analyses. For intra-CTM, fused rawdata prediction has a higher optimization rate than the error rate which shows a similar gain trend, and about 80% of the optimized patients appear in at least 2 feature extraction networks. For inter-CTMs, eighty-five percent of the patients that all CTMs predicted incorrectly have optimizable RGMs within each CTM. The results also proved that the gain of rawdata is stable across different convolutional networks and different CTM approaches. Therefore, exploring the drawbacks of post-reconstructed CT image analysis and developing models for direct diagnosis from rawdata are the keys to future research. The results of the subgroup analysis showed that the RGMs performed better than the CTM in most subgroups, especially in the subgroup of older, female, and smaller lesion size, indicating that the rawdata could provide more valuable information that brings model gains in subgroups, while this information may have been lost in the process of CT reconstruction.

Our study has some limitations. First, this study involved a small number of patients and the proportion of positive and negative samples is unbalanced. Further study on large-scale multicenter datasets should be performed. Second, only patients with single nodule are included in this study, further validation of our method on patients with multiple nodules should be further studied. Third, although the rawdata had a comparable performance with CT, it still had a certain gap with the best CT diagnosis. There is an urgent need to develop novel AI methods specifically for rawdata.

Meanwhile, strong computing power is a problem that cannot be ignored when calculating rawdata. It is not realistic to read the complete high-frequency scanning data directly to the computing device. Designing appropriate pre -processing algorithms and building deep networks in combination with characteristics of rawdata are the potential breakthrough points in the future. Finally, the CT scan scheme is designed for image reconstruction and it may be not suitable for rawdata analysis. Therefore, novel scan strategies, e.g., scanning for specific diagnostic purposes, should be developed to maximize the gain of rawdata.

In conclusion, for the first time, we validated the potential value of rawdata in real clinical practice. The rawdata analysis showed comparable performance with CT images, which indicates that leveraging non-image information holds promise as an alternative to image-based methods. Moreover, the added value of rawdata to CT images was also confirmed in this study, which means that the combination of non-image and image will further promote the advance of disease diagnosis. This study proposed and validated a new feasible direction for diagnosis without image reconstruction, and it may facilitate the development of fully automated scanning and diagnostic processes.

## Methods

### Patients

In this prospective study, 626 consecutive patients who had a chest CT scan in the First Hospital of Jilin University from November 2019 to May 2021 were recruited. Eligible patients were included according to the following inclusion criteria: (i) patients who had a pulmonary lesion more than 2 cm with contrast enhanced chest CT scan, (ii) rawdata obtained from CT machine after the imaging examination, (iii)pathological diagnosis of pulmonary lesion with two weeks interval from CT scan. Patients were excluded on the basis of the following: (i) previous systemic antineoplastic treatments, (ii) CT images with poor image quality or unreadable scan. After exclusion, a total of 276 patients were included for modeling experiments.

The methods were performed in accordance with Standards for Reporting Diagnostic accuracy studies (STARD) and approved by the Ethics Committee of the First Hospital of Jilin University (AF-IRB-032-05).

### Collection of CT image and rawdata

Both the CT images and rawdata were consecutively collected from the First Hospital of Ji Lin University and were acquired with a NeuViz Prime CT system (Neusoft Medical Systems Co., Ltd., Shenyang, China). The system parameters of CT scanner included source-to-isocenter distance of 570 mm, source-to-detector distance of 1040mm, and scanning FOV of 500 mm. The imaging protocol included contrast-enhanced CT of the chest with variable imaging parameters. Contrast-enhanced CT scans were performed at a spiral scan mode using 324 mA tube current, 100 kVp tube voltage, 0.5 s ration time, and 0.9 spiral pitch. CT rawdata were reconstructed using a kernel F20 at slice thickness of 1.0 mm with image pixel range from 0.59 mm to 0.98 mm and image matrix of 512 by 512. In addition, we also acquired the initial height and initial view angle of the CT detector each time the patient underwent a scan. Finally, CT images and rawdata from each scanner were randomly stratified into one of three cohorts in a 6:2:2 ratio: a training cohort, a validation cohort and a test cohort. All in all, **Table S4** describes CT scanner information, system parameters and imaging parameters.

### lesion segmentations in CT images

The segmentations of primary lesion were manually delineated across all the sections in the axial view using annotation tool in ISD (IntelliSpace Discovery, Philips, German). The regions of interest were annotated and reviewed by four radiologists with 8 to 25 years’ chest CT experience. All radiologists were blinded to any clinical or histopathologic information. The annotation was labeled as five common categories according lesions’ pulmonary lobe.

### Realization of typical CT models

There are many studies on benign-malignant lung nodule classification on chest CT. We selected four typical papers from the major journals, including *IEEE Transactions on medical imaging, Medical image analysis*, and *Nature medicine*, which refer to multi-scale ensemble method (CTM1) [27], global and local information fusion method (CTM2) [26], loss function-based method (CTM3) [28], and multi-view fusion (CTM4) method [29]. We further performed experiments on four typical models with our dataset, and all the realization details are described in **Supplementary 1**.

### Extraction of lesion region from raw data

After acquiring 4 CTMs, we proceeded to perform rawdata gain experiments. The first step of the experiment is to select projection surface containing lesions in the rawdata. The rawdata of CT scans contains three dimensions: 1) the index dimension representing the acquisition order; 2) the projection surface which is the detector receives the x-ray attenuation, where the channel and row directions are defined as x and y, respectively. All lesion segmentation regions of rawdata were derived from the binarized segmentation of CT image after being represented in a unified coordinate system. The complete derivation can be condensed into three steps: orientation, querying and mapping.

#### 1) Orientation

On the derivation of localization, we took the segmented regions in the CT image as the research object, and the localization calculation mainly includes cross-sectional localization and height localization. For cross-sectional positioning, we set the center point between the CT source and detector as the coordinate origin (which is also the rotation center of the CT gantry), parallel to the cross-section of the CT image. Next, the motion trajectory can be characterized by the scan index *t*, the rotation radius *r* and the angle *θ*. In order to obtain the above parameters, we first read the origin coordinates (*x*_origin_, *y*_origin_ and *z*_origin_), and the offset values (*x*_offset_ and *y*_offset_) can be calculated through voxel spacing and image size (*x*_size_ and *y*_size_).

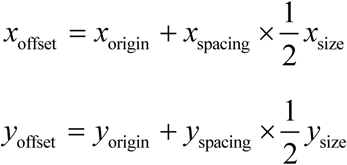

Next, with the help of the offset values and the voxel coordinates *x*_CT_ and *y*_CT_ in the CT image, the distance from the coordinate origin (*x*_length_ and *y*_length_) can be calculated as:

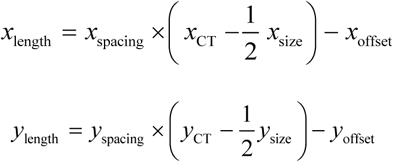

Otherwise, the radius *r*, and the starting angle *θ*_0_ can be obtained.

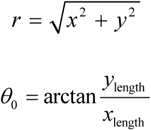

Meanwhile, by introducing the scanning period of the machine, we got the angle change Δ*θ* with the following relationship:

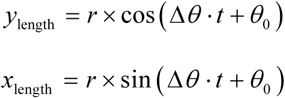

For height positioning, we directly obtained the initial height *h*_start_ of the voxel through the coordinate *z*, slice thickness and origin of the voxel point in CT images.

#### 2) Querying

Our purpose in this step is to determine the interval of index dimension *t* in which tumor voxel appears in the raw data. Since there is a cone beam in the projection, we first calculated the change function *h*_area_ of the voxel, where *d* is the distance from the voxel to the X-ray focal spot on the x-axis; *l* is the distance from the focal spot to the detector. The number of detector rows is *n*_*y*_ ; the channel spacing along y-axis is Δ*n*_*y*_.

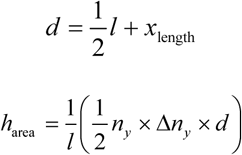

Next, we determined the index (*t*) range of the voxel in the rawdata by the following inequality, where *h*_0_ is the initial height at which the detector start to scan; Δ*h* is the height change in a scan.

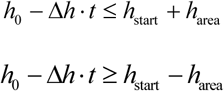

To reduce computational complexity, we first extracted the highest and lowest masks in the segmentation images, and calculated the start and end indices of two voxels. Then, we initially located the range of index dimensions. Within this interval, we computed the mapping result of voxels within the layer.

#### 3) Mapping

Through the above calculation, we have obtained the index interval corresponding to the voxel, then the voxel appearing in index is obtained by calculating the projection data of the index layer by layer. The coordinates of each voxel on the projection surface are defined as *x*_raw_ and *y*_raw_, respectively. *Y*_raw_ is related to the height *h*_t_, *h*_area_ at the *t* index and the number of detector rows *n*_y_ in the detector, so we determined its height difference relative to the detector by the following formula, and then calculated its coordinates in the projection.

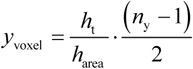

Since the x-axis of the projection plane is equiangularly sampled, *x*_raw_ can be acquired through the angle at the *t* index *θ*_t_, the view angle *θ*_d_ of the detector, and the number of channels in the detector *n*_x_.

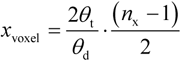

After obtaining the segmentation files of lesions in the raw data, we saved the raw data segment through the initial index interval, and used this as the training data for this gain experiment. It should be added that there are different directions in the actual retrieval of raw data (From head to foot or foot to head). We used the same spatial relationship to modify the inequality for different directions and then located the lesion.

### Construction of RGM based on CT images

To explore whether the rawdata contained unique information, we built residual fusion models through the rawdata and fused it with CTMs’ output to determine whether the rawdata could bring benefits. First, we built three feature extraction networks using the rawdata. Based on the memory need of calculation, we sampled the index dimension of rawdata fragments containing lesions to one-eighth, and the same size was resampling based on the average value by equal interval sampling. For the Channel dimension, we directly removed the data outside the reconstruction area from both sides, and resampling with the row dimension into half of the size. For model building, we did not modify Densenet121 [21], Resnet18 [22] and Resnext18 [23] in 3D with the purpose of directing the direct gain of nude data as much as possible. The training settings and parameters are detailed in **Supplementary 2**.

The core of the residual fusion model is to obtain the correction of the CT model output, and the origin of the idea is that the learning residual is easier which is mentioned in Resnet. The probabilities of predicting the patients as positive by the CTMs were fused with the predicted probabilities of the rawdata models. This fusion is performed during the training process. Specifically, the probability of predicting one patient as positive was calculated as:

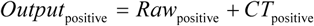

The probability of predicting one patient as negative was calculated as:

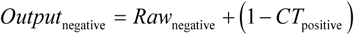

After the output fusion of the CTM and rawdata model, the loss function was used to calculate the loss and optimize the model. The three feature extraction networks built with rawdata were fused with the four representative CTMs described above to obtain four raw gain models respectively, which were: raw gain model-Densenet121 (RGM-DN 1/2/3/4), raw gain model-Resnet18 (RGM-RE 1/2/3/4), raw gain model-Resnext18 (RGM-RX 1/2/3/4). Therefore, we obtain 12 raw gain models. Then the RGMs were compared with the CTMs to evaluate the benefits of the rawdata.

### The calculation of the average attention score

For calculating the average attention score of each voxel, we first used the segmentation data of lesion in rawdata to obtain non-lesion area by unary complement. Next, we dotted and summed the segmentation data of lesion and non-lesion areas with the attention matrix. Finally, the average attention score was obtained by dividing the total amount of attention in the two areas by the number of voxels in the segmented regions, respectively. It should be noted that we also normalized the average attention score of the lesion area and the non-lesion area in each rawdata, so as to obtain a more intuitive comparison result.

## Data Availability

The image features and output values associated with the CTMs and the RGMs are stored on GitHub (https://github.com/CASIAMI/rawdata_gain). The original data that support the findings of this study are available from the corresponding author upon reasonable request.

https://github.com/CASIAMI/rawdata_gai

## Code availability

Source code for related CT and raw data methods can be found from GitHub (https://github.com/CASIAMI/rawdata_gain).

## Acknowledgements

This work was supported by the National Key R&D Program of China (2017YFA0205200), National Natural Science Foundation of China (82022036, 91959130, 81971776, 62027901, 81930053, 81771924), the Beijing Natural Science Foundation (Z20J00105), Strategic Priority Research Program of Chinese Academy of Sciences (XDB38040200), Chinese Academy of Sciences under Grant No. GJJSTD20170004 and QYZDJ-SSW-JSC005, the Project of High-Level Talents Team Introduction in Zhuhai City (Zhuhai HLHPTP201703), the Youth Innovation Promotion Association CAS (Y2021049) and the China Postdoctoral Science Foundation (2021M700341). The authors would like to acknowledge the instrumental and technical support of Multi-modal biomedical imaging experimental platform, Institute of Automation, Chinese Academy of Sciences.

## Author contributions

B.X.H., Y.B.Z., C.X.S., T.T., K.S. and H.L.L. developed the network architecture and data/modeling infrastructure, training and testing setup. B.X.H., Y.B.Z., C.X.S., T.T., K.S. and H.L.L. wrote the methods. B.X.H., C.X.S. and T.T. created the figures. B.X.H. and M.J.F. performed statistical analysis. J.T., D.D., Z.Y.L., K.W., Z.Q.W., W.M., S.W., Z.C.T., S.T.Z., J.W.W. and L.Z.S. advised on the modeling techniques. L.X.T., L.W.W., S.P.Z. and J.T.L. provided raw data structure information. D.D., B.X.H., Y.B.Z., C.X.S., T.T., K.S. H.L.L., L.W.W. and Y.G. wrote the manuscript. H.M.Z., Y.G., M.W., F.Y.M., L.D., L.L.Z. and S.W. provided clinical expertise and guidance on the study design. H.M.Z., Y.G., M.W., F.Y.M. and L.D. created the clinical datasets, interpreted the data and defined the clinical labels. L.X.T., W.L.W., and F.H. created the rawdata sets. J.T., D.D., F.H. and H.M.Z. initiated the project and provided guidance on the concept and design. J.T., F.H. and H.M.Z. supervised the project.

## Figures and Tables

**Extended Data Fig. 1.**
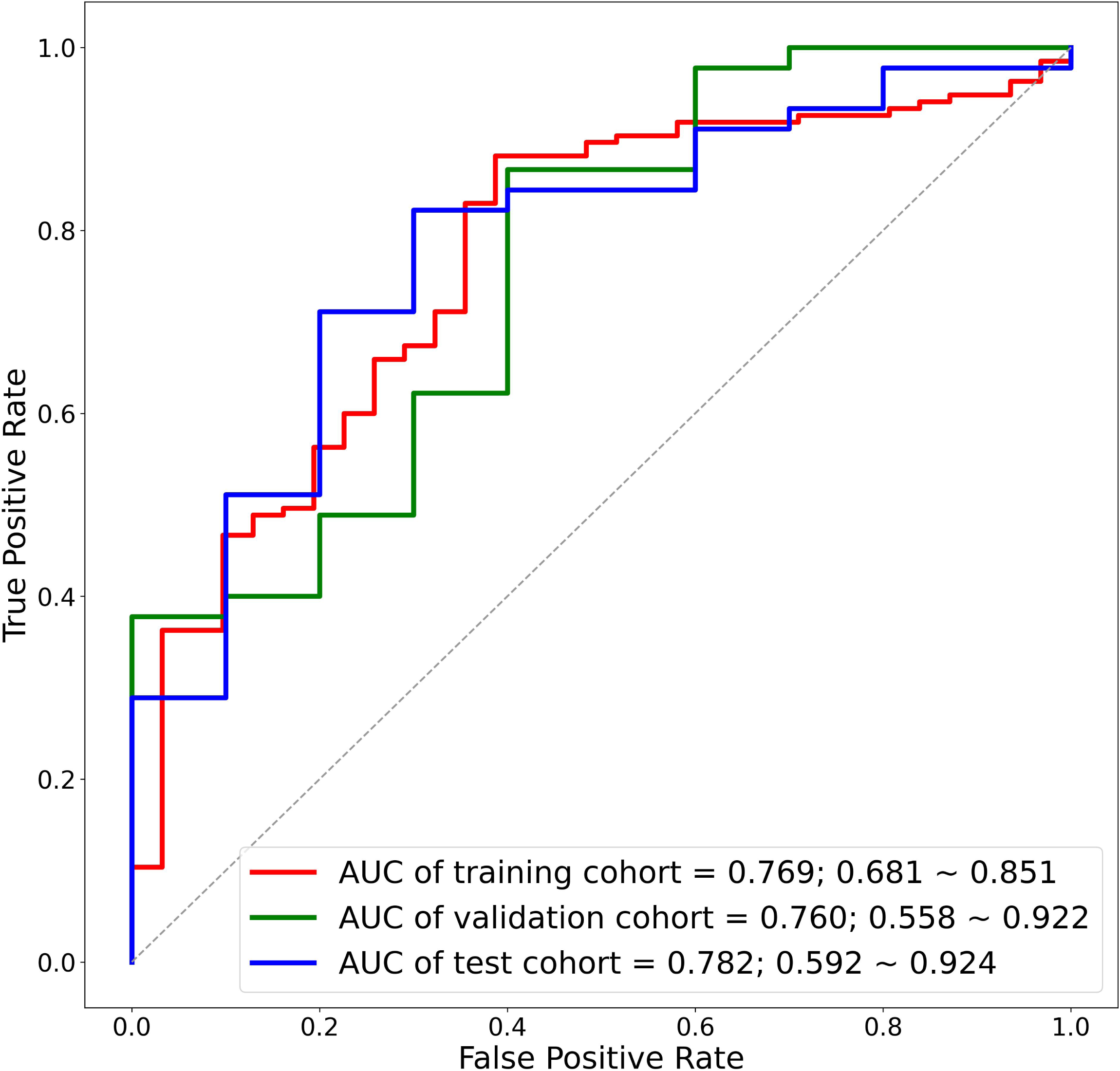
The ROC curve of the model built by rawdata only.

**Table S1.**
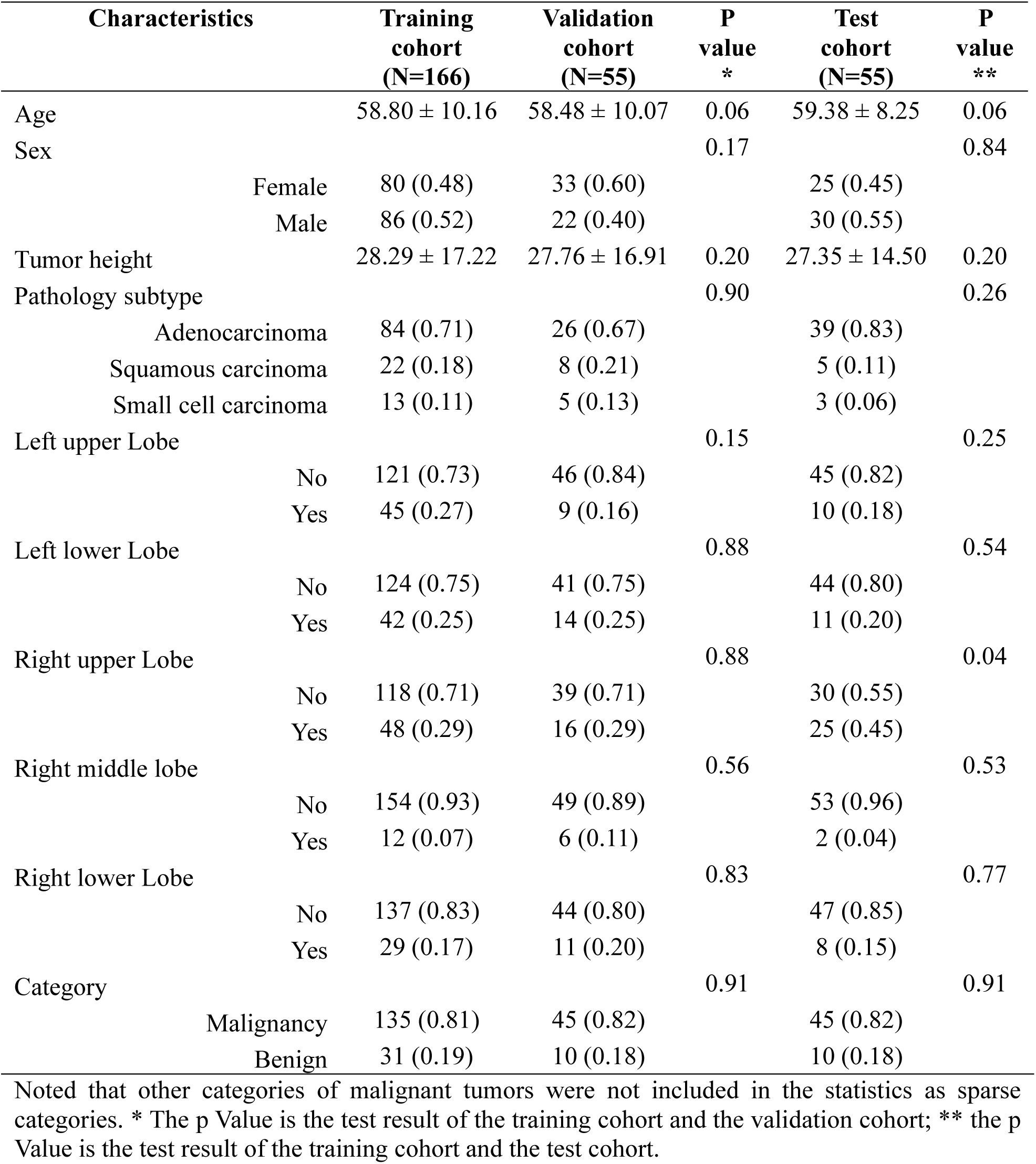
Clinical characteristic statistics

**Table S2.** Detailed gain statistics of raw data for CT models

| ID of CT model | Optimization rate of error sample | Error rate of correct sample | Ratio of at least 2 gain model optimizations |
| --- | --- | --- | --- |
| CTM1 | 0.771 (81/105) | 0.329 (57/173) | 0.827 (67/81) |
| CTM2 | 0.676 (46/68) | 0.286 (60/210) | 0.848 (39/46) |
| CTM3 | 0.724 (76/105) | 0.208 (36/173) | 0.789 (60/76) |
| CTM4 | 0.406 (26/64) | 0.079 (17/214) | 0.846 (22/26) |

**Table S3.** The performance of four CT models in subgroup analysis.

| AUC<br>95%CI |  | CTM 1 | CTM 2 | CTM 3 | CTM 4 |
| --- | --- | --- | --- | --- | --- |
| Age | ≤60 | 0.790<br>(0.707-0.861) | 0.739<br>(0.626-0.831) | 0.801<br>(0.729-0.866) | **0.831**<br>**(0.749-0.904)** |
|  | >60 | 0.705<br>(0.546-0.823) | 0.725<br>(0.592-0.849) | 0.701<br>(0.599-0.799) | **0.790**<br>**(0.672-0.899)** |
| Sex | Male | 0.765<br>(0.680-0.848) | 0.748<br>(0.641-0.866) | 0.721<br>(0.626-0.803) | **0.804**<br>**(0.706-0.882)** |
|  | Female | 0.752<br>(0.638-0.858) | 0.729<br>(0.626-0.827) | 0.803<br>(0.703-0.888) | **0.823**<br>**(0.720-0.920)** |
| Tumor<br>Size | ≤23mm | 0.760<br>(0.647-0.843) | 0.696<br>(0.569-0.809) | 0.753<br>(0.664-0.833) | **0.806**<br>**(0.699-0.888)** |
|  | >23mm | 0.764<br>(0.670-0.852) | 0.798<br>(0.697-0.890) | 0.774<br>(0.684-0.862) | **0.819**<br>**(0.719-0.906)** |
| Tumor<br>Location | superior<br>left | 0.802<br>(0.671-0.901) | 0.671<br>(0.463-0.880) | 0.873<br>(0.749-0.961) | **0.921**<br>**(0.843-0.980)** |
|  | inferior<br>left | 0.698<br>(0.526-0.852) | 0.732<br>(0.557-0.868) | 0.698<br>(0.562-0.840) | **0.812**<br>**(0.675-0.934)** |
|  | superior<br>right | 0.759<br>(0.615-0.878) | 0.732<br>(0.578-0.852) | 0.731<br>(0.603-0.844) | **0.807**<br>**(0.683-0.912)** |
|  | Inferior<br>right | 0.781<br>(0.630-0.906) | **0.843**<br>**(0.712-0.950)** | 0.769<br>(0.617-0.901) | 0.695<br>(0.443-0.928) |

**Table S4.** Scanner information, system parameters and imaging parameters for the Chest CT examinations

| Manufacturer | Scanner | System parameters | Contrast-enhanced CT imaging parameters |
| --- | --- | --- | --- |
| Neusoft Medical Systems Co., Ltd., Shenyang, China | NeuViz Prime | Source-to-detector distance, 1040 mm;<br>Source-to-isocenter distance, 570 mm;<br>Scanning FOV, 500 mm;<br>Scanning frequency, 2320 times/round;<br>Detector channel, 672 pcs;<br>Detector row, 64 pcs;<br>Detector channel spacing, 0.625 mm. | Tube current, 324mA;<br>Tube voltage, 100kV;<br>Rotation time, 0.5 s;<br>Spiral pitch, 0.9;<br>Image matrix, 512x512;<br>Pixel spacing, 0.59~0.94 mm<br>Slice thickness, 1mm;<br>Kernel, F20;<br>Image time relative to onset of contrast material injection, pre-contrast 60s~70s. |

## Supplement 1. Implementation and optimization details of typical CT models

Considering the differences in experimental design between our study and the typical models, we need to make appropriate modifications to make the typical models have the best performances on our dataset. The difference is mainly reflected in data volume and data imbalance. The amount of data in previous articles ranged from 1018 to more than 20,000, much larger than that in this experiment (276). Therefore, we applied small data learning related techniques (including pre-trained model and sharing network weight) for four typical CT models. Hence, in our study, a pre-trained 3D-resnet18 was used for 3D input, which was pre-trained using eight medical datasets [30], a pre-trained 2D-resnet50 was used for 2D input. To train the subnets effectively using our small dataset, only the last layer (layer 4) and full connected network were trained in this study. In addition, for data imbalance, we used resampling strategy during training.

Specific to each of these models, we describe them in detail below. CTM1 was a multi-scale ensemble model, which ensembled three subnets whose networks were all the same, except for the input size. The input data were cropped from CT images using three different sizes, 32×32×32, 48×48×48, and 64×64×64 pixels. The outputs of three subnets were weighted to obtain the final ensemble results, and a grid search was used to tune the weight values. In addition, AUC loss proposed in original paper was used for dealing with data imbalance. CTM2 combined the features of both the entire CT volume and the region of interest cropped from CT volume, so that all predictions relied on both nodule-level local information and global context from the entire CT volume. In addition, CTM2 was trained with focal loss to mitigate the data imbalance. CTM3 designed a deep network with a margin ranking loss to enhance the discrimination capability on ambiguous nodule cases. In our study, 3D input was used to preserve the spatial information of pulmonary nodule. CTM4 was a multi-view deep network, which ensembled the outputs of nine 2D-view subnets to characterize the 3D nodule. Each subnet combined three types of image patches, and each patch was input into a pre-trained ResNet-50 network. In this study, all networks shared weights. Finally, nine subnets were used jointly to classify nodules with an average weighting scheme. For model training, the batch size was set as 32. The cross-entropy loss was used as the loss function, and the SGD optimizer was applied. The start learning rates were set as 0.001, and the models were trained for 100 epochs.

## Supplement 2. Model training and statistical analysis methods

For model building, the batch size was set as 64. The Cross-Entropy loss was used as the loss function, and the Adam optimizer was applied. The start learning rates were set as 1 × 10^−5^, 1 × 10^−6^ separately, and the learning rate decay was set as 0.001. Meanwhile, we also added weight attenuation. The weight decay was set as 0.01, 0.05, and 0.1 separately. The model was trained for 50 epoches and select the best parameters with the lowest loss of validation cohort. The three RGMs built based on the three structures were all trained with the above setting.

For statistical analysis, discrete variables and continuous variables are calculated in the chi-squared test and Man-Whitney U test, respectively. All models are implemented in Python 3.7.3 (https://www.python.org/) with Numpy (version ≥ 1.16.4), SciPy (version ≥ 1.3.0), Matplotlib (version ≥ 3.1.1), Scikit-Learn (version ≥ 1.10.1), Statsmodels (version ≥ 0.12.2) and Pandas (version ≥ 0.25.0). All models were trained in python package named Pytorch (version ≥ 1.10.1; https://pytorch.org/) with 4 Graphics Processing Units of NVIDIA TITAN RTX (24G).

